# The impact of human behavioral adaptation stratified by immune status on COVID-19 spread with application to South Korea

**DOI:** 10.1101/2024.11.19.24317549

**Authors:** Sileshi Sintayehu Sharbayta, Youngji Jo, Jaehun Jung, Bruno Buonomo

## Abstract

As the COVID-19 pandemic continues with ongoing variant waves and vaccination efforts, population-level immunity and public risk perceptions have shifted. This study presents a behavioral transmission model to assess how virus spread and care-seeking behavior differ based on individuals’ immunity status. We categorized the population into two groups: “partially immune” and “susceptible,” which influenced their response to vaccination and testing, as well as their prioritization of information related to disease prevalence and severity. Using COVID-19 data from South Korea (February 1, 2022 - May 31, 2022), we calibrated our model to explore these dynamics. Simulation results suggest that increasing reactivity to information among partially immune individuals to the same level as susceptible individuals could reduce peak active cases by 16%. Conversely, if partially immune individuals shift their risk perception focus from prevalence (90% prevalence vs. 10% severity) to severity (90% severity vs. 10% prevalence), the peak in active cases could increase by 50%. These findings highlight the need for adaptive vaccination and testing strategies as public risk perceptions evolve due to prior exposures and vaccinations. As new variant waves emerge in the post-pandemic endemic era, our study offers insights into how immunity-based behavioral differences can shape future infection peaks.

**Subject class:** 92D30, 92-10, 37N25, 34A34

## 1 Introduction

COVID-19 increasingly appears likely to enter long-term circulation and become endemic, necessitating regular vaccinations with updated vaccines, similar to seasonal influenza[1, 2]. Changes in risk perceptions during the pandemic affect behaviors, including testing and willingness to vaccinate [3]. Patterns of COVID-19 transmission shape the subsequent patterns of behavioral responses to the disease and, in turn, are shaped by such responses. Some models have been developed to help policymakers compare interventions such as testing and vaccination [4, 5]. Such models, dependent on various uncertain assumptions, attempt to forecast cases, deaths, and medical supply needs; predict the timing of peaks in cases; and estimate if and when to expect additional waves or surges. Despite the rapid advancements in COVID-19 models and forecast tools, very few models [6, 7, 8] directly incorporate adaptive behavioral components to account for changes in risk perceptions, protective behaviors, and compliance with interventions over time, which ultimately influence transmission.

Compliance with testing and willingness to vaccinate significantly affect disease transmission dynamics and can influence policy recommendations. Hence, modeling how voluntary testing compliance and vaccination willingness are influenced by individual immune status/history and available public information (e.g., rumors on vaccine efficacy or level of prevalence/severity) is crucial. Coupling transmission models of infectious diseases with models of behavior adaptation has been a growing field of research [9, 10, 11]. More specifically, an information index approach was recently employed in epidemic models to account for the social behavior change due to available information related to the disease status in the population [6, 8, 12, 13, 14]. This strategy takes into consideration how the information is distributed; the dependence of human behavior not only on the current knowledge but also on the past state of the disease in the population; and how long it takes for the information to reach the general public (information delay). For example, vaccination behavior change due to available information about the prevalence has been considered in a meningitis model[13]. In [14], the authors considered a general SISV (SusceptibleInfectious-Susceptible-Vaccinated) model where transmission (social distancing compliance behavior) and vaccination rate (vaccination decision) depend on prevalence and vaccination roll-out, respectively. Most of these studies, however, often assume a homogeneous population where all individuals react similarly to a single type of information, such as disease prevalence, without considering other critical factors like immune status, severity, mortality rates, or healthcare system strain.

In this study, we develop a compartmental model to represent the transmission dynamics of COVID-19 structured by two different susceptible populations (naive vs. partially immune due to previous infection or vaccination). We then incorporate the information index approach, where the individuals’ compliance of vaccination and testing is based on two different kinds of information, namely information about the level of prevalence and severity of the disease. We also take into account the change of risk perception by differing the weighting on information between prevalence and severity among people who are partially immune and not immune (i.e., susceptible) [15, 16, 17].

The paper is organized as follows: we present the model formulation (Section 2), basic properties of the model (Section 3), and model fitting and parameter estimation (Section 4). We then conduct various numerical simulations (Section 5), which show the role of information-related parameters on the dynamics of the disease. Finally, we discuss the model findings and implications (Section 6).

## 2 Model formulation

The entire population (N) is divided into fifteen distinct compartments according to individuals’ infection and vaccination status. We divide the transmission dynamics into two categories: primary dynamics, which describes disease transmission for susceptible individuals, and secondary dynamics, which describes transmission among partially immune individuals due to vaccination or previous infection. Each dynamic’s transmission is discussed below.

### (i) Primary and secondary dynamics

In primary dynamics there are six compartments: susceptible (*i*.*e*., non-immune) individuals (*S*_1_), those who are not yet infected and are susceptible to infection; Vaccinated (*V*_1_), individuals who got vaccinated with primary series vaccination; Exposed (*E*_1_), individuals who are infected but not yet infectious; Asymptomatic (*A*_1_), people who are infected but do not show symptoms of the disease; Symptomatic people (*I*_1_), those who are infected and show symptoms of the disease; Tested (*I*_*T* 1_), individuals who get tested to COVID-19 and their result is positive. Individuals in *S*_1_ class get infected at a rate of *λ*_1_ (the force of infection) and join the exposed class. People in the *E*_1_ class either join the *A*_1_ class at the rate of (1 *−τ*)*ϵ* or *I*_1_ class at the rate of *τϵ*, where *τ* represents the proportion of exposed individuals who become symptomatic and *ϵ*^*−*1^ is the latency period. Individuals in the *A*_1_ and *I*_1_ classes get tested for COVID-19 at the rate of *ξT*_1_ and *T*_1_ respectively and join the *I*_*T* 1_ class, where *T*_1_ is the testing rate. Due to the illness differences among individuals in *A*_1_ and *I*_1_ classes, we assumed their test seeking rates will be different. Therefore, *ξ* measures the propensity for testing of individuals in the asymptomatic class relative to the symptomatic class. In secondary dynamics, there are seven compartments. These are: susceptible (partially immune) compartments, *S*_2_ and *S*_3_, that contain people who have vaccination history and who have vaccination or infection history respectively, Booster-vaccinated, (*V*_2_); Exposed (*E*_2_), these are individuals infected but not infectious; Asymptomatic (*A*_2_), individuals who are infectious but have no symptoms; Symptomatic (*I*_2_), infectious with symptoms; Tested (*I*_*T* 2_), who are tested for COVID-19 and their result is positive. It has been demonstrated that the immunity level acquired from prior infection, primary series vaccination, and a combination of both (hybrid immunity) vary in their effectiveness against secondary infection or hospitalization [18, 19]. Our study considers the immunity effectiveness remaining until six months after the infection/vaccination based on the study [18], which we utilize to determine protection levels against infection for immune people. Consequently, individuals in the *S*_2_, *S*_3_, and *V*_2_ classes get infected at the reduced rates of (1 *− η*_2_)*λ*_2_, (1 *− η*_3_)*λ*_2_ and (1 *− η*_4_)*λ*_2_, respectively, due to these differing immunity levels. The parameters *η*_2_ and *η*_3_ measure the effectiveness of the immunity gained due to vaccination and prior infection, respectively, after 6 months, whereas *η*_4_ represents the effectiveness of booster vaccination. However, people who get the primary series vaccination, *V*_1_, can get infected at the rate of (1 *−η*_1_)*λ*_2_, where *η*_1_ measures the effectiveness of the first series vaccination. The transition from primary dynamics to secondary dynamics is either from *V*_1_ or recovery, *R* at the rate of *ϕ*. People in the *S*_2_ and *S*_3_ classes get vaccinated at a rate of *F*_2_, this represents the booster vaccination. People in the *E*_2_ class either join the *A*_2_ class at the rate of (1 *− τ*)*ϵ* or the *I*_2_ class at the rate of *τϵ*, where *τ, ϵ* are as explained in primary dynamics. Individuals in *A*_2_ and *I*_2_ classes get tested for COVID-19 at the rate of *ξT*_2_ and *T*_2_, respectively, and join the *I*_*T* 2_ class, where *T*_2_ is the testing rate and *ξ* measures the lower propensity for testing of individuals in the *A*_2_ compared to the *I*_2_ class. People in *A*_1_, *A*_2_, *I*_1_, and *I*_2_ classes recover from the disease at the rate of *ρ* (assumed to be equal), and people in *I*_*T* 1_, *I*_*T* 2_ recover at the rate of *ρ*_*t*_ and join *R*, class. Individuals in *I*_*T* 1_ and *I*_*T* 2_ classes may progress and be hospitalized, joining the *H* compartment at the rate of *h*_1_ and *h*_2_ respectively. Vaccination against COVID-19 helps prevent severe illness, resulting in a lower hospitalization rate among individuals in the secondary dynamics (*h*_2_ *< h*_1_). Hospitalized individuals recover from the disease at the rate of *ρ*_*h*_. Individuals in *I*_1_, *I*_2_, and *H* classes die due to COVID-19 at rates *d*_1_ and *d*_2_ respectively. People are recruited into the Susceptible, *S*_1_, class by birth at a rate of *π*, and individuals in all compartments die naturally at rate *µ*. The schematic diagram of the model is shown in Figure 1. The infection, vaccination and testing rate formulations are explained in depth in the following sub-sections:

**Figure 1:**
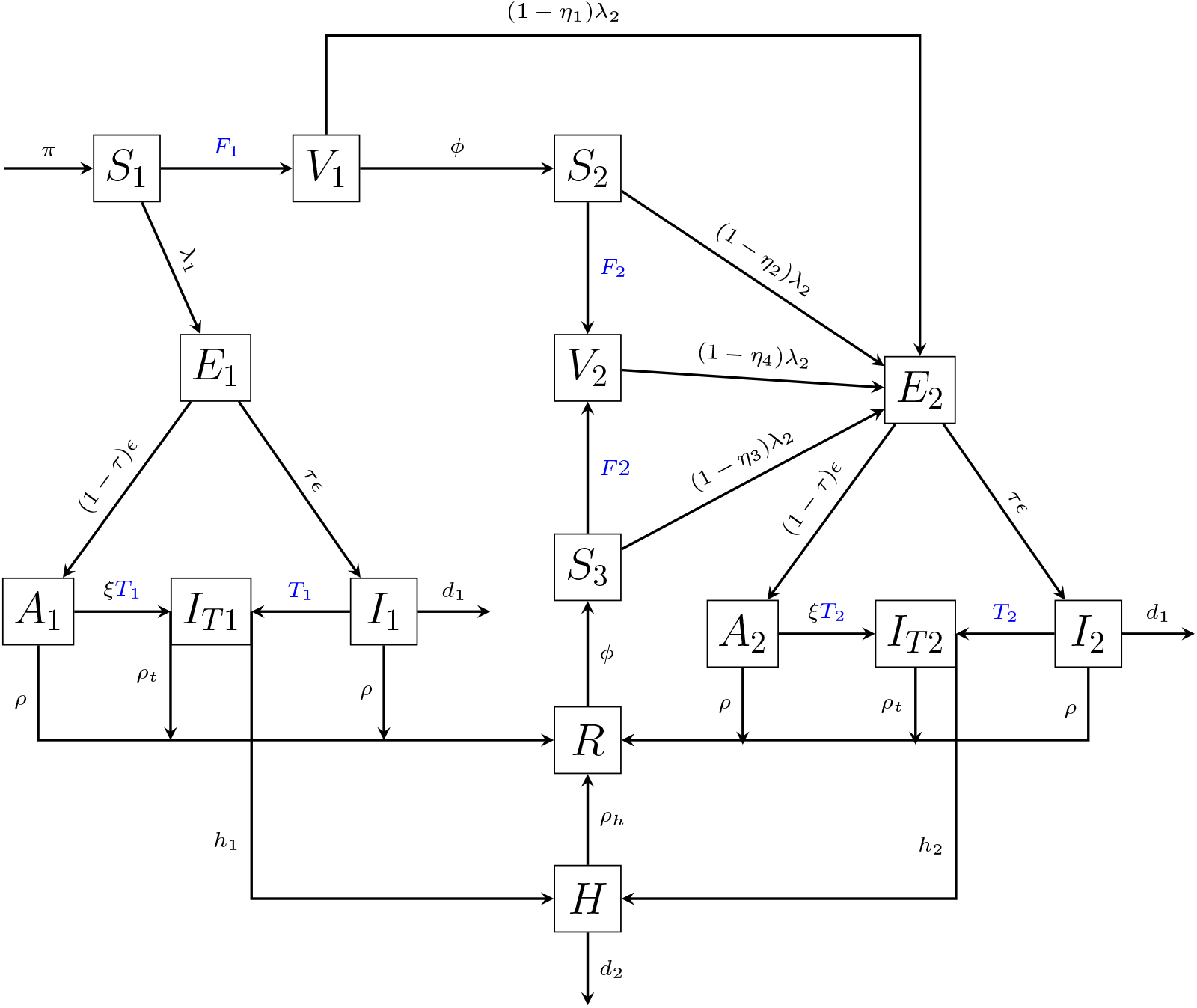
Transmission dynamics of COVID-19. The blue-colored parameters are information dependent parameters. *F*_1_ is a primary series vaccination rate and *F*_2_ is a booster vaccination rate. *T*_1_ and *T*_2_ are testing rates in primary and secondary dynamics respectively.

### (ii) Force of infection

Due to immune differences, the transmission rate in primary and secondary dynamics is different [20, 21]. Given that mandatory quarantine measures are not commonly enforced nowadays, we assume that a certain proportion of individuals who test positive and all hospital admissions will opt to self-quarantine or isolate themselves, thus they don’t contribute to the transmission [22]. With these assumptions, the forces of infection are given by:

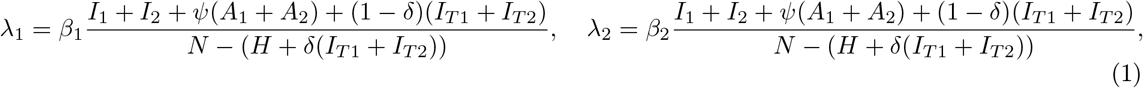

where *N* is the total population and is given by :

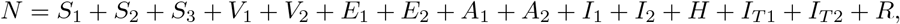

and *β*_1_ and *β*_2_ are the transmission rates in primary and secondary dynamics respectively, *δ* is percentage of tested individuals who quarantine and *ψ* represents infectiousness modification of asymptomatic individuals.

### (iii) Vaccination and testing rates

The vaccination and testing rates are both described by the sum of two rates: mandatory and voluntary rates. First, the mandatory vaccination and testing rates represent the rates for the portion of the population that will be vaccinated or tested regardless of the information. This term summarizes some aspects of vaccine acceptance or test seeking of individuals that are strongly in favor of vaccines or advised to get tested, or specific population groups (e.g., older age groups, teachers or health workers) for which the vaccination or testing is mandatory or strongly recommended by the authorities. These rates are represented by a constant rate. These constant rates are represented by *F*_10_ (mandatory primary series vaccination rate), *F*_20_ (mandatory booster vaccination rate) and *T*_10_ (mandatory testing rate in primary dynamics), *T*_20_ (mandatory testing rate in secondary dynamics), respectively. Second, the voluntary rate is a rate for a portion of the population voluntarily choosing to be vaccinated or tested depending on the level of disease prevalence and severity in the society. We use the information index to represent the publicly available information or rumors about the prevalence and severity of the disease. We use the reported number of people dead and hospitalized to represent the level of severity of the disease. In these formulations, we made two assumptions: first, partially immune people have a lower perception of the risk of infection, and second, partially immune and susceptible people prioritize prevalence and severity information differently. The Holling type *II* function, (characterized by saturating, continuous, differentiable, and increasing function), is commonly used to represent the voluntary rate [8, 13]. Based on the above discussion, the vaccination and testing rates are given by:

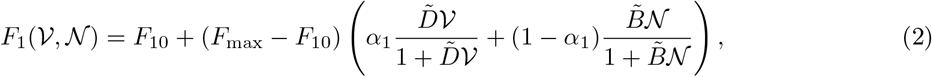

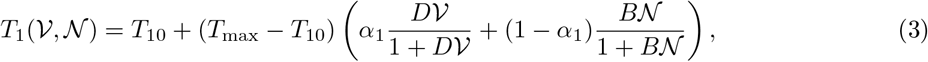

and

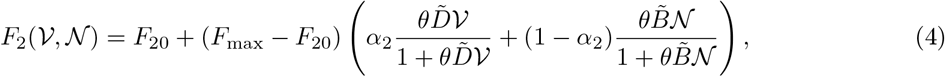

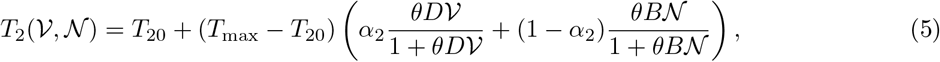

where 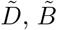, *D*, and *B* are positive factors that adjust the reactivity of individuals to the information in vaccination (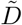, and 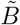) and testing (*D* and *B*) rates, *F*_max_ and *T*_max_ represent the maximum vaccination and testing rates, respectively, that can be achieved in the case of a high level of information coverage about the disease status (or high level of risk perception), *θ* represents the reduced reactivity to risk perception by individuals in the second dynamics relative to individuals in primary dynamics. The weight given by individuals to the prevalence information in primary and secondary dynamics is measured by *α*_1_ and *α*_2_, respectively. The corresponding complementary weights, 1 *− α*_1_ and 1 *− α*_2_, are assigned to the severity information. The variables *V* and *N* are the information indices that represents the information available to the public or rumors about the level of prevalence and severity, respectively and are given by the following delayed distributions:

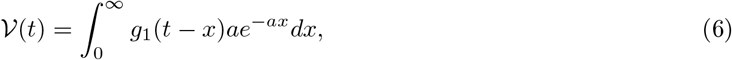

and

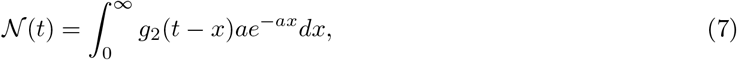

where *a* represents the inverse of the average time delay for the information to reach the general public, and the functions *g*_1_(*t*) and *g*_2_(*t*) represent people’s perception of the risk of infection and the level of severity of the disease, respectively (often called message functions). Perceived risk of infection and severity are assumed to depend on the number of symptomatic individuals (*I*_1_ + *I*_2_ + *I*_*T* 1_ + *I*_*T* 2_ + *H*) and number of hospitalized and dead people (*H* + *d*_1_(*I*_1_ + *I*_2_) + *d*_2_*H*) respectively. Thus, the message functions are given by

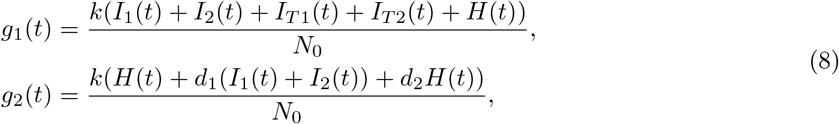

where the quantity *k* represents information coverage. We define information coverage as the publicly available information or rumors about the disease status [13, 14] and *N*_0_ is the steady state population when there is no disease and no disease-induced death (*i*.*e*., 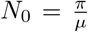). Asymptomatic people are often unreported and hidden from the public, therefore they are not included in information indices. The formulation in equations (6) and (7) represents that the population’s memory about the perceived risk is fading exponentially. The term *ae*^*−at*^ is often known as the *exponential fading kernel* [23]. It represents the weight given to the current and past values of the disease. Utilizing the *linear chain trick* method [24], the integral equations (6) and (7) can be reduced into ordinary differential equations (ODEs), given by

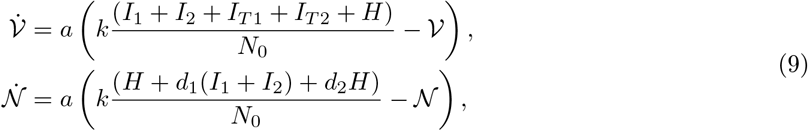

where the upper dot denotes the time derivative.

Based on the above discussions, the system under study is governed by the following non-linear ODEs.

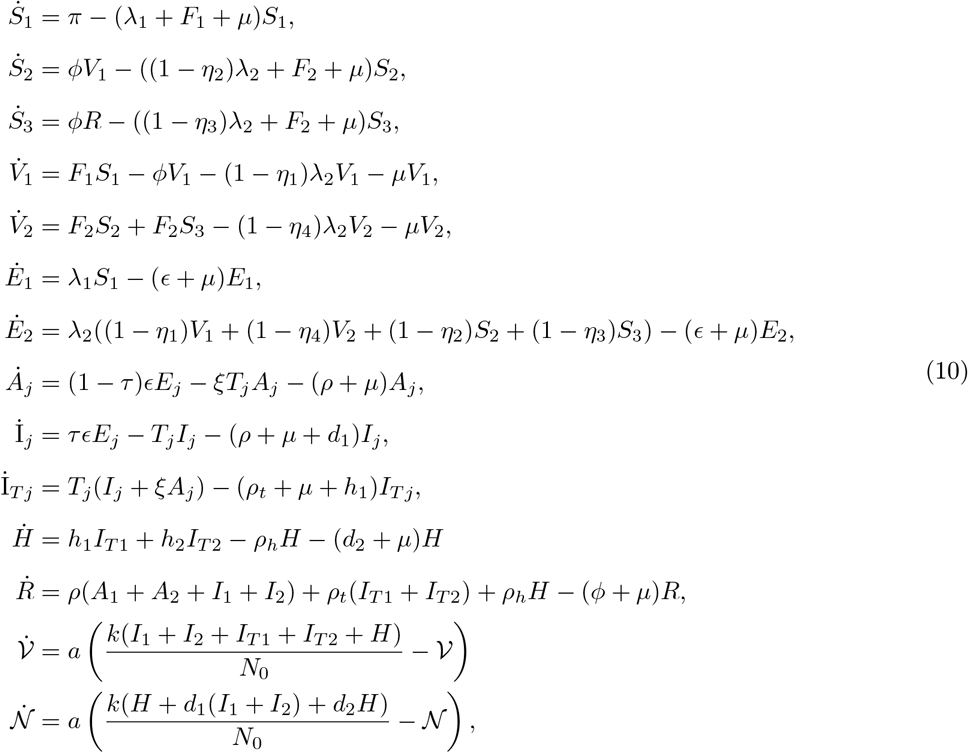

with initial conditions:

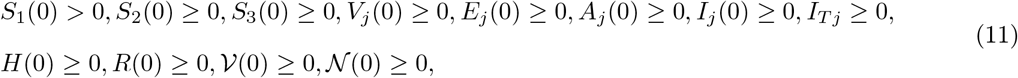

where *j* ∈ *{*1, 2*}*.

## 3 Basic properties

In this section, we investigate the basic characteristics of the model (10), which include ensuring the positivity and boundedness of solutions, computing the disease-free equilibrium, and determining the reproduction number.

### (i) Positivity and boundedness of solutions

Positivity and boundedness of the solutions can be established in standard ways (see e.g., [25, 26]).

### (ii) Disease-free equilibrium and the reproduction number

A disease-free equilibrium point is an equilibrium point at which there are no infected individuals in the population (no disease). Setting all infected compartments of the system (10) to zero and solving the reduced system of equations by equating to zero, we get the disease-free equilibrium point, denoted by *E*^0^, and given by

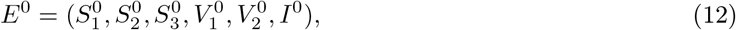

where

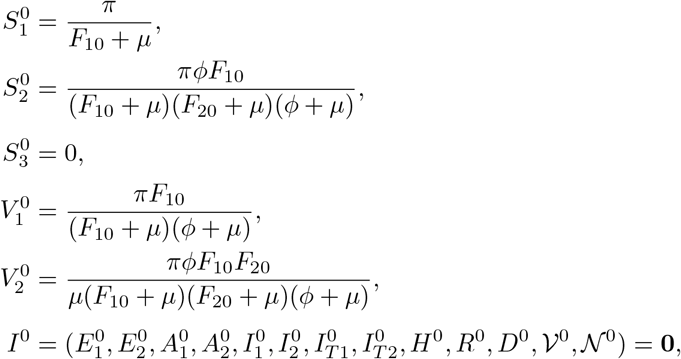

where **0** represents a zero vector of dimension 1 *×* 13.

The effective reproduction number is the expected number of new infections caused by an infectious individual in a population where some individuals may no longer be susceptible (due to obtained immunity from prior infection or vaccination) [27].

We used the next-generation matrix method to calculate the effective reproduction number for the model (10). This method follows the following three steps (for detailed explanations, one can refer to [28, 29]):

*Step I:* We sort out the equations for infected compartments (*E*_1_, *E*_2_, *A*_1_, *A*_2_, *I*_1_, *I*_2_, *I*_*T* 1_, *I*_*T* 2_, *H*) and split the right-hand side of the equations as

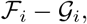

where ℱ_*i*_ represents the rate of appearance of new infections in compartment *i* and 𝒢_*i*_ incorporates the remaining terms representing the transition of people into and out of the compartments.

*Step II:* Determine the following matrices that are obtained by linearizing the equations in Step I and evaluating at the disease-free equilibrium.

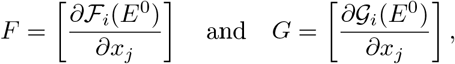

where *x* represents the infected compartments.

The next-generation matrix is defined as

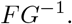

*Step III:* Find the reproduction number using

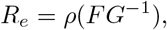

where *ρ* is the spectral radius of the matrix and is defined as the maximum of the absolute values of the eigenvalues of the matrix *FG*^*−*1^.

Following the above steps, the effective reproduction number, *R*_*e*_, is given by

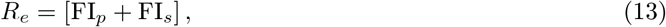

where

FI_*p*_ and FI_*s*_ represents the effective reproduction number for primary dynamics and secondary dynamics, respectively, and are given by

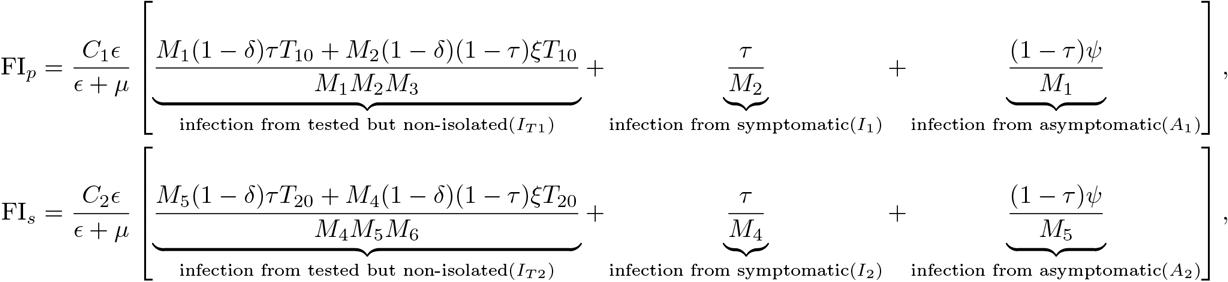

and,

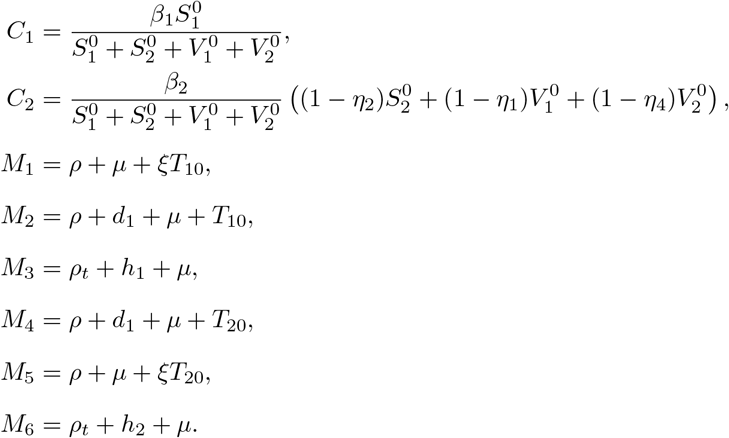

#### Remark 1. Note that

- *The basic reproduction number, i*.*e*., *the mean number of secondary cases a typical single infected case will cause in a fully susceptible population in the absence of interventions, for the model* (10) *can be found by setting the mandatory vaccination rates (F*_1_0 *and F*_20_*) and mandatory testing rates (T*_10_ *and T*_10_*) to zero*.
- *The parameters related to voluntary vaccination and testing (like, information coverage, information delay time, reactivity to information, information prioritization and reduced level of reactivity by immune individuals) don’t appear in the effective reproduction formula. Hence, they will not affect the threshold value (R*_*e*_ = 1*) in Theorem 1*.

**Theorem 1**. *The disease-free equilibrium, E*^0^, *of the model* (10) *is locally asymptotically stable if R*_*e*_ *<* 1 *and unstable if R*_*e*_ *>* 1.

*Proof*. The proof follows from Theorem 2 in [29].

## 4 Model fitting and parameter estimation

In this section, we will discuss how we set the model’s baseline parameter values. The parameter values in the model are determined in two ways: first, using demographic and epidemiological data obtained from Our World in Data [30] and previous research, which is mentioned as a reference in Table 1, and second, by fitting the model to the Korean COVID-19 vaccination, incidence and mortality data collected during the omicron variant wave with time period from February 01, 2022, to May 31, 2022. We will discuss the details of each method in the following sections.

**Table 1:** Parameters with their description and baseline values.

| Param. | Description | Baseline value | Reference |
| --- | --- | --- | --- |
| $\pi$ | Recruitment rate to susceptible class | 1710 indivi./day | See Sec. 4(*i*) |
| $\mu$ | Natural death rate | $3.301 \times 10^{-5}$ | See Sec. 4(*i*) |
| $\epsilon$ | Reciprocal of latency period | 0.33 | [35] |
| $\psi$ | Reduced infection rate for asymptomatic class | 0.5 | [36, 31] |
| $\tau$ | Proportion of individual that become symptomatic | 0.8 | [32] |
| $\rho$ | Recovery rate for symptomatic & asymptomatic class | 1/14 | [35] |
| $\rho_t$ | Recovery rate for tested class | 1/10 | Assumed<br>($\rho_t > \rho$) |
| $\rho_h$ | Recovery rate for hospitalized class | 1/10 | Assumed<br>($\rho_t = \rho_h$) |
| $\eta_1$ | Primary series vaccine effectiveness | 0.71 | [19] |
| $\eta_2$ | Primary series vaccine effectiveness after 6 months | 0.41 | [18] |
| $\eta_3$ | Booster Vaccine effectiveness | 0.85 | [37] |
| $\eta_4$ | Effectiveness of immunity due to prior infection after 6 months | 0.46 | [18] |
| $\phi$ | Transition from Recovery and primary vaccination classes to secondary dynamics | 1/180 | [18] |
| $h_1$ | Hospitalization for tested class (primary dynamics) | 0.0012 | [38] |
| $h_2$ | Hospitalization for tested class (secondary dynamics) | 0.000312 | [18] |
| $F_{\max}$ | Maximum vaccination rate | 0.027 | See Sec. 4(*i*) |
| $F_{10}$ | Mandatory vaccination in primary dynamics | 0.0029 | See Sec. 4(i) |
| $F_{20}$ | Mandatory vaccination in secondary dynamics | 0.00261 | See Sec. 4(i) |
| $T_{\max}$ | Maximum testing rate | 0.5 | See Sec. 4(i) |
| $T_{10}$ | Mandatory testing in primary dynamics | 0.03 | See Sec. 4(i) |
| $T_{20}$ | Mandatory testing in secondary dynamics | 0.03 | See Sec. 4(i) |
| $a$ | Inverse of the average information delay time | 0.33 | Assumed |
| $\tilde{D}$ | Reactivity factor to voluntary testing (primary dynamics) | 5 | See Sec. 4(i) |
| $\tilde{B}$ | Reactivity factor to voluntary vaccination (primary dynamics) | 5 | See Sec. 4(i) |
| $B$ | Reactivity factor to voluntary testing (secondary dynamics) | 5 | Assumed $B = \tilde{B}$ |
| $D$ | Reactivity factor to voluntary vaccination (secondary dynamics) | 5 | Assumed $D = \tilde{D}$ |
| $\theta$ | Reduced reactivity to the information in secondary dynamics | 0.5 | Assumed |
| $\delta$ | Percentage of positively tested people who quarantine | 80% | Assumed |
| $\alpha_i, i = 1, 2$ | Weight given to the information | 0.5 | Assumed |
| $d_1$ | Disease-induced death rate for symptomatic class | 0.000071 | Iteratively chosen |
| $d_2$ | Disease-induced death rate for hospitalized class | 0.000073 | Iteratively chosen |
| $\beta_1$ | Transmission rate (primary dynamics) | 0.67 | Fitted |
| $k$ | Information coverage | 0.5046 | Fitted |
| $\beta_2$ | Transmission rate (secondary dynamics) | 0.39 | Fitted |
| $\xi$ | Testing modification for asymptomatic class | 0.99 | Fitted |

### (i) Values of known parameters and initial conditions for the model

According to the data from Our World in Data, the estimated population of South Korea in 2022 is *N* (0) = 51815808 (the initial population used in the simulation) and the life expectancy is 83 years [30]. Therefore, the daily natural death rate can be calculated as 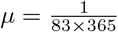 and the daily birth rate is obtained by *π* = *µ × N* (0) = 1710 [31]. The mandatory primary series vaccination rates and the maximum vaccination rate are estimated from the data [30]. We estimated the maximum vaccination rate, *F*_max_, to be the maximum proportion of daily vaccinated people given by: 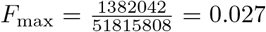. The mandatory vaccination rate, *F*_10_, is obtained by calculating the average of the daily proportion of vaccinated people prior to the initial time for our simulation (February 01, 2022), under the assumption that this value can represent the baseline vaccination rate (not influenced by the current level of omicron prevalence or severity) before the omicron wave. Thus, we found *F*_10_ = 0.0029. We assumed a lower (10%) rate for mandatory booster vaccination rate, *F*_20_ = 0.9 * 0.0029 = 0.00261, compared to the mandatory primary series vaccination rate. We iteratively increased the mandatory testing rates, *T*_10_ and *T*_20_ starting from 0.0025 (average proportion of reported daily tested people prior to February 01, 2022), with the intention of including the rate for people who can undergo testing at home, to achieve a best fit to the data.The process resulted in *T*_10_ = *T*_20_ = 0.03. Assuming substantial number of people can get tested (both at home and health centers) we fixed the value of *T*_max_ to be 0.5. At the beginning of a vaccination campaign, the cumulative number of vaccinated individuals is assumed to grow linearly. The values for reactivity factors in vaccination, 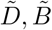 (in primary dynamics, in secondary dynamics) are chosen so that the vaccination rate can represent an initial linear growth. We accomplished such a choice by iteratively plotting the vaccination rate for different randomly chosen values of 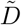 and 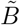 in the interval [1, 20]. The bounds of the interval are arbitrarily set to include a reactivity value (2.5) used in the study [14]. Thus, we found 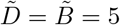. We assumed a similar pattern for reactivity parameters in testing rates, so that *D* = *B* = 5. The initial conditions are fixed by the data as follows: as of Feb 01, 2022, we assumed 85% of the total population (44, 365, 186 out of 51, 815, 808) gained immunity from prior infection/vaccination and were partially susceptible with waning immunity over time. Therefore, they were initially assigned to *S*_2_ or *S*_3_ classes. Assuming a few of them get immunity through infection only [32], we distributed them as about 90% are in *S*_2_ and 10% in *S*_3_. Therefore, *S*_2_(0) = 40, 000, 000, *S*_3_(0) = 4, 365, 186. Based on the initial population distribution (85% in secondary dynamics and 15% primary dynamics), we used the same distribution for the initial conditions for infected and vaccinated compartments in both dynamics. For example, from the data, the number of new vaccinated people on February 01, 2022 was 3, 323. Therefore, 85% of these people are in *V*_2_ (most of this is booster vaccination), and the remaining are in *V*_1_. That is, *V*_1_(0) = 431 and *V*_2_(0) = 2891. Similarly, the initial conditions for tested compartments, which are daily new cases, became *I*_*T* 1_(0) = 2634 and *I*_*T* 2_(0) = 17632. According to the study [33] it was assumed that the number of exposed cases is 20 times the number of symptomatic cases. Therefore, the total number of exposed people is assumed to be 405, 339, which is distributed as *E*_1_(0) = 52, 694 and *E*_2_(0) = 352, 645. According to the surveillance data of South Korea [32] 80% of exposed people become symptomatic and 20% become asymptomatic. Therefore, we set *A*_1_(0) = 0.2 * *E*_1_(0) = 10, 538, *I*_1_(0) = 0.8 * *E*_1_(0) = 42, 155 and *A*_2_(0) = 0.2 * *E*_2_(0) = 70529, *I*_2_(0) = 0.8 * *E*_2_(0) = 282, 116. From Our World in Data, we used the number people in the ICU on February 01, 2022 as an estimate for the initial population in the hospital; therefore, *H*(0) = 203. The initial condition for the death compartment is, *D*(0) = 15, that is, the number of death on February 01, 2022. We assumed recovered individuals to be *R*(0) = 200, more than 10 times the number of deaths. This reflects that many more people recover from the disease after experiencing mild illness due to their immunity. Finally, the rest of the population is placed in the *S*_1_ compartment. *i*.*e. S*_1_(0) = *N* (0) *− Σ*_*i*_ *Z*_*i*_(0), where *Z*_*i*_ represents all compartments in the model except *S*_1_ and *D*. As for the information indices, we set their initial conditions at the equilibrium [14]: *i*.*e*., *V*(0) = *k*(*I*_1_(0) + *I*_2_(0) + *I*_*T* 1_(0) + *I*_*T* 2_(0) + *H*(0))*/N*_0_, *N* (0) = *k*(*d*_1_(*I*_1_(0) + *I*_2_(0)) + *d*_2_*H*(0) + *H*(0))*/N*_0_.

### (ii) Model fitting with South Korea COVID-19 data

We fitted the model (10) to the cumulative daily cases, daily vaccination, and daily death data for the time period from February 01, 2022 to May 31, 2022. We fit our model and conducted numerical simulations to the highest peak of the omicron wave to evaluate how intervention scenarios and information parameters can influence the peak of the epidemic. The fitting process is accomplished by a Python built-in curve fitting function called *curve_fit* (Nonlinear least squares optimization method) [34]. In general, this method identifies the best parameter values by minimizing the sum of squared errors between the model output and the data sets. For our model, there are four parameters to be estimated: transmission rates in primary and secondary dynamics (*β*_1_ and *β*_2_), information coverage (*k*), and testing modification of asymptomatic individuals (*ξ*).

The result of the fitting is displayed in Figure 2. The model best approximates the cumulative daily cases (panel a) and slightly overestimates the death data (panel c). Although, the final cumulative death data approximation is close to the observed data. The cumulative vaccination data follows some irregular patterns as compared to the daily cases and death data. The model approximation to the cumulative vaccination data changes over time (it slightly underestimates,or overestimates, panel b). The parameter descriptions and their baseline values are displayed in Table 1.

**Figure 2:**
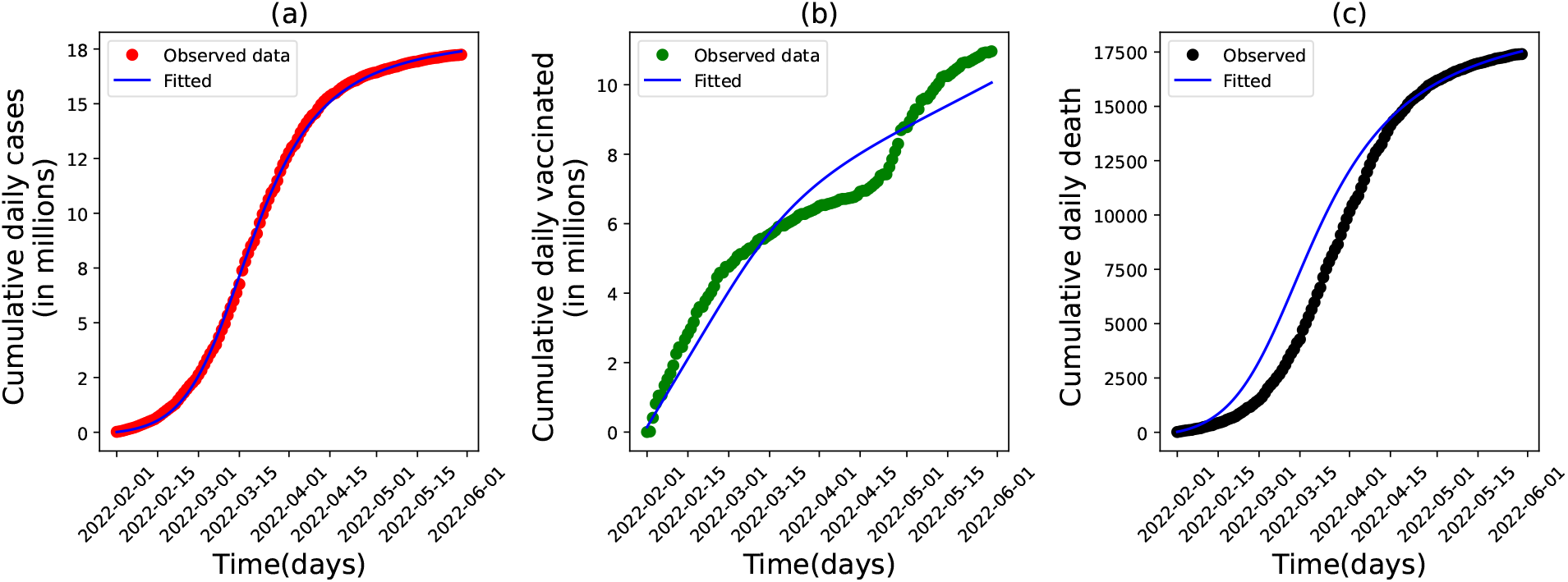
Result of the model fitting to South Korea’s COVID-19 epidemic data. The data represents the cumulative observed daily cases (in panel a), vaccination (in panel b) and death (in panel c). The time frame is from February 01, 2022 to May 31, 2022. The blue curve shows the approximation by the model (10). The approximation for a number of cases is obtained by adding a number of individuals in *I*_*T* 1_ and *I*_*T* 2_ classes (who are tested positive). Similarly, the estimated number of vaccinations is obtained by summing the number of individuals in *V*_1_ and *V*_2_ classes. The initial conditions used for daily cases, daily vaccination and daily death are *V*_1_(0) + *V*_2_(0) = 3322, *I*_*T* 1_(0) + *I*_*T* 1_(0) = 20266 and *D*(0) = 15, respectively.

In Figure 3, we displayed the time series of daily cases of observed data (green scattered points), the approximation by the model when voluntary vaccination and testing is considered (purple curve) and not considered (blue curve). The latter case is obtained by setting the reactivity parameter values to zero 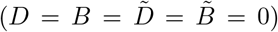. We call a result of the model a *responsive case (base case)*, when the vaccination and testing rates are ruled by both mandatory and voluntary rates (people’s vaccination choice is affected by the level of the disease status) and *unresponsive cases* when these rates are ruled only by the mandatory rates (level of prevalence or severity does not affect the individual’s vaccination and testing decision). The result in Figure 3 shows that the model best estimates the observed daily cases when voluntary vaccination and testing are considered. When the voluntary part is not considered, the model underestimates the number of daily cases. For instance, the peak of active is lower (by 33%) in the unresponsive case compared to the base case (responsive case) (*i*.*e*., a peak decrease from 366280 to 246389).

**Figure 3:**
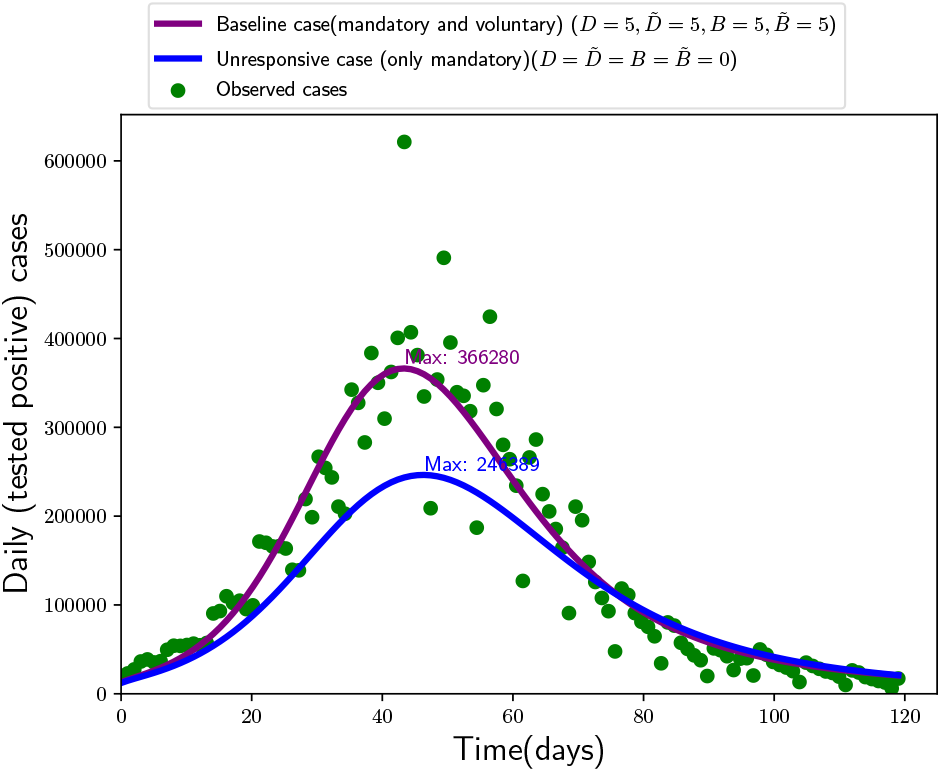
A comparison of observed daily cases (green scattered plot) with model approximation of daily cases when the vaccination and testing rates are ruled by both mandatory and voluntary rates (purple curve) and when these rates are ruled by only mandatory rate (blue curve).

With the parameter values in Table 1, the estimated value of the effective reproduction number is *R*_*e*_ = 0.64. This value is the sum of the effective reproduction number in primary dynamics and secondary dynamics, being 0.07 and 0.57, respectively. The basic reproduction number is estimated to be 8.43. This value falls in the interval [5.5, 24], which contains the basic reproduction values for omicron virus that are reported in different studies [39]. The effective reproduction number we found is smaller than the values reported in some other studies [40, 41]. In [40], the mean reproduction number for the omicron variant during the first local outbreak in South Korea was estimated to be 1.72. Another study [41] estimated the effective reproduction number for the omicron variant as 1.3 during the time period November 25, 2021 – January 08, 2022. A review paper [39] showed that the effective reproduction number for omicron reported in different studies fall between 0.88 and 9.4. The variation in the reproduction number across different studies can be attributed to several factors, such as the time and locations of the studies, exposure patterns, vaccine coverage, and the levels of immunity in the affected populations. In our model, a key reason for the low effective reproduction number is the initial population distribution, where 85% of the population is initially immune, with vaccine efficacy ranging from 41% for the primary series vaccination to 85% for booster vaccination. For instance, if the booster vaccination efficacy decreases to 75% (10% lower than the base case, 85%), the effective reproduction number increases to 0.9989.

### (iii) Stability region for the disease-free equilibrium in (F_10_, F_20_) plane

From a vaccination and testing perspective, the parameters that drive the effective reproduction number are those related to the mandatory vaccination and testing rates (*F*_10_, *F*_20_, *T*_10_, *T*_20_). Thus, by varying these rates, one can achieve the condition *R*_*e*_ *<* 1 (stability of the disease-free equilibrium). Since varying the mandatory testing rates, *T*_10_ and *T*_20_, does not modify *R*_*e*_ beyond one, we vary the mandatory vaccination rates in primary dynamics (*F*_10_) and secondary dynamics (*F*_20_) here to depict their impact on the stability region. The result is shown in Figure 4 where the shaded region is the region in which *R*_*e*_ *>* 1 (showing instability of the disease-free equilibrium) and the non-shaded region is where *R*_*e*_ *<* 1 (showing stability of the disease-free equilibrium). One can set threshold values for these parameters, given that the other parameter values in the model are kept constant. For example, when the first mandatory vaccination rate - *F*_10_ in the primary dynamics is less than 0.0005 or the mandatory booster vaccination rate - *F*_20_ is less than 0.00016, then *R*_*e*_ *>* 1 regardless of the wide variation of the primary series or boosting vaccination rate, respectively. However, if *F*_10_ *>* 0.00075 and *F*_20_ *>* 0.0002, then the disease spread can be controlled over time (*R*_*e*_ *<* 1). The boundary of the regions (*R*_*e*_ = 1) in the figure is where the stability of the disease-free equilibrium changes.

**Figure 4:**
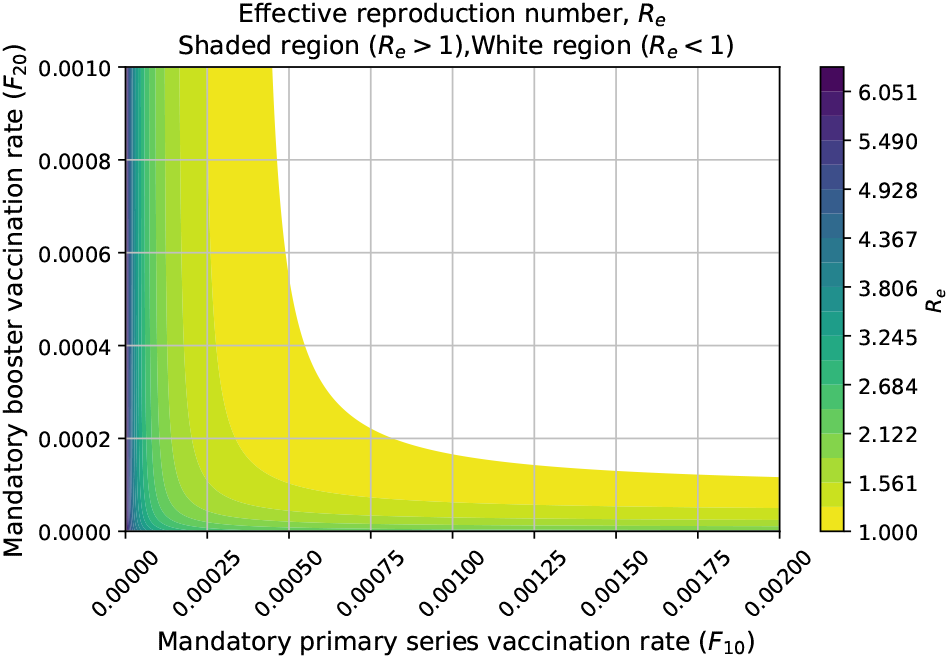
Value of the effective reproduction number *R*_*e*_ by varying the mandatory primary series vaccination rate, *F*_10_ and mandatory boosting vaccination rate, *F*_20_. The shaded region is a region where *R*_*e*_ *>* 1 and the white region is where *R*_*e*_ *<* 1. Other parameter values are fixed to their baseline values, as in Table 1.

## 5 The role of behavior-related parameters

In this section, we perform numerical simulations to investigate the effect of parameters related to voluntary vaccination and testing on the disease dynamics. Except the varying parameters in the plots, all other parameter values are fixed as in the Table 1.

### (i) Impact of changes of information parameters on active cases peak

Here, we address the effect of changes in information coverage, information delay and reactivity factors on active cases peak. Active cases indicate the number of infectious individuals who are not detected (tested), those are in *A*_1_, *A*_2_, *I*_1_ and *I*_2_. Figure 5 illustrates that information coverage is the key driver among other parameters. The maximum active cases is more sensitive to the information delay when the information coverage is larger than the base line value (*>* 51%). The minimum peak of active cases is attained when there is a higher information coverage (*k >* 80%) and shorter time delay (*T* ≤ 3 days), see Figure 5 panel (a). For example, when *k* = 0.9 and *T* = 3 days the peak of active cases is around 4.3 million, where as when *k* = 0.1 and *T* = 30 days the peak is almost double as 8.2 million. In Figure 5 panels (b) and (c) we assumed reactivity to prevalence in vaccination, 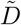, and testing, *D* to vary equally. A similar assumption is made for reactivity to severity parameters: in vaccination 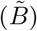 and testing (*B*). The results in Figure 5 panels (b) and (c), show that the peak of active cases is more sensitive to an individual’s reactivity to information about prevalence than to information about severity. When people react to the prevalence, 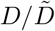 is less than 5 (base value); increasing information coverage does not affect the active cases peak, Figure 5 panel (b). This demonstrates that prompt public response is an essential factor in addition to the government’s efforts to achieve high information coverage in a timely way to reduce the epidemic peak.

**Figure 5:**
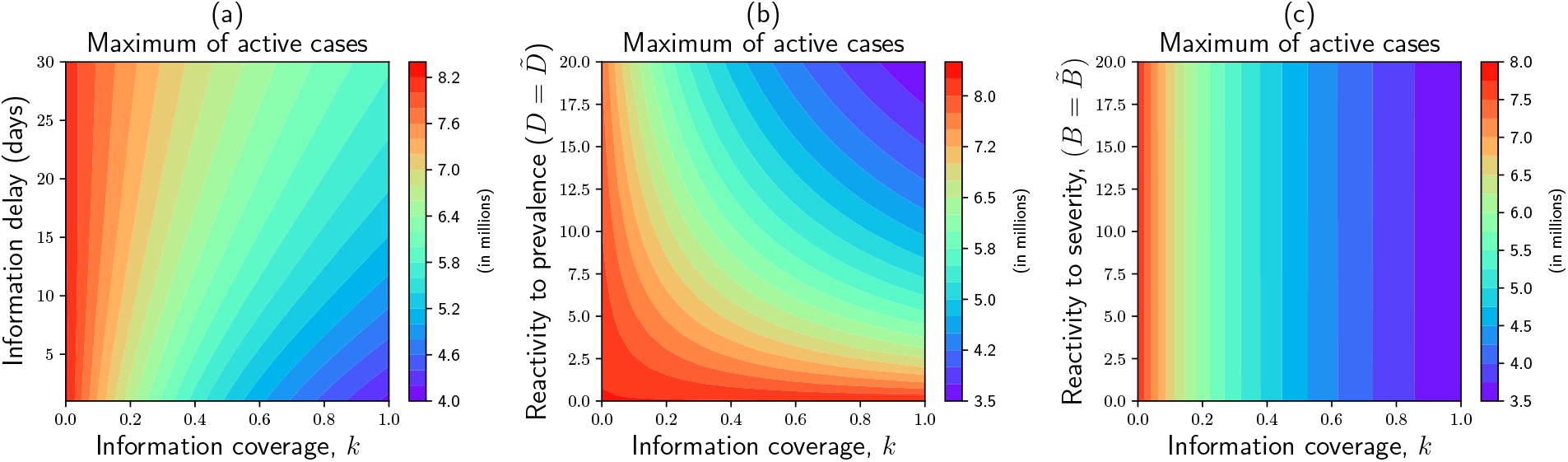
Contour plot for maximum (peak) of active cases by varying information coverage (*k*) with: information delay time (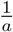 days), panel (a), people’s reactivity to prevalence information (assumed to vary equally, 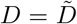, in vaccination and testing rates), panel (b), reactivity to severity information (assumed to vary equally, 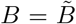, in vaccination and testing rates), panel (c). Active cases represent a number of infectious individuals who are undetected (not tested) (*A*_1_ + *A*_2_ + *I*_1_ + *I*_2_).

### (ii) Impact of change in level of reactivity to the information by partially immune population on the dynamics of active cases

Here, we will explore how the reduction in reactivity to information among partially immune individuals, *θ*, compared to susceptible ones, impacts the dynamics of the disease, particularly the number of active cases. At its base line value, *θ* = 0.5 (50% reduction), the active cases peak becomes 5.5 million, we call this as the base active case peak. The lowest active cases peak is achieved when partially immune people react to information the same level as the susceptible ones (*θ* = 1), which is 16% less than the base active cases peak and 40% less than the active cases peak when partially immune people not reacting to the information (*θ* = 0), see Figure 6. When *θ* = 0.75 (25% reactivity reduction by immune people compared to the susceptible individuals), the peak decreases by 9% compared to the base case. In general, the active cases peak lowers as partially immune people behave more similarly to non-immune persons (*i*.*e*., *θ* changes from 0 to 1).

**Figure 6:**
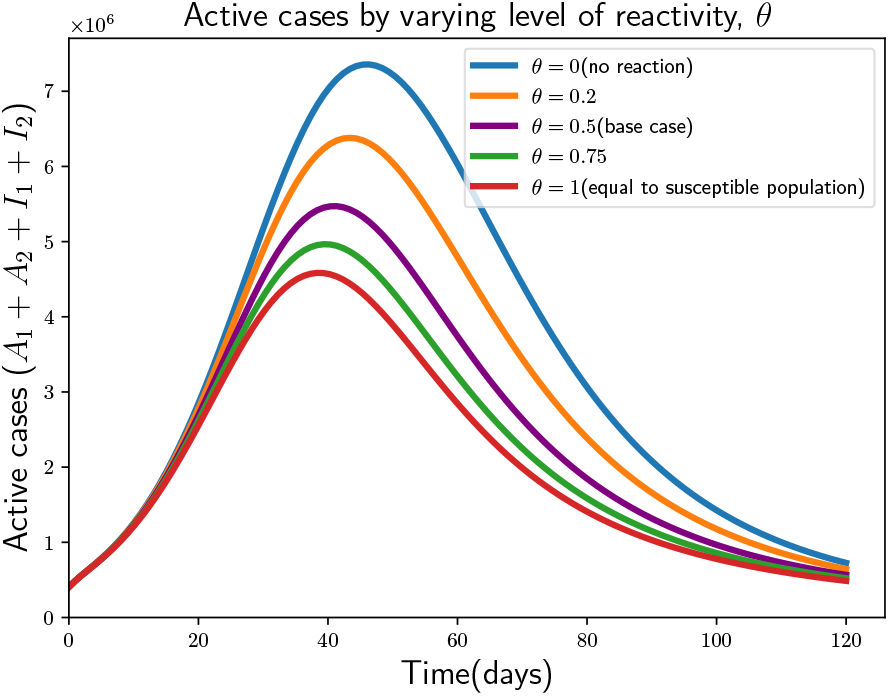
Time series of active cases, with varying levels of reactivity to the information by partially immune people(people in secondary dynamics) relative to susceptible (people in primary dynamics). The baseline value for the level of reactivity to information by susceptible individuals is 5.

### (iii) Impact of behavior adaptation between immune and susceptible populations on dynamics of vaccination and testing rates, and active cases

The compliance of individuals with protective measures may vary over the course of a pandemic [16, 17], especially in scenarios where the disease persists despite widespread vaccination, as seen in the case of COVID-19. In such circumstances, it becomes essential to analyze the dynamics by accounting for behavioral disparities among sub-populations and their different risk perceptions. We accounted for the disease status information (prevalence and severity) that can influence the sub-populations’ (susceptible and partially immune) risk perception and compliance with vaccination and testing (i.e., susceptible people may prioritize prevalence while partially immune people may prioritize the severity (representing a change in risk perception [17]), and this phenomenon will impact their compliance to vaccination and testing differently). We assessed the consequence of vaccination and testing rates and the epidemic burden (number of active cases) by applying different weights (using *α*_1_ and *α*_2_ for prevalence and 1 *− α*_1_ and 1 *−α*_2_ for severity, respectively) between two information (prevalence and severity) among the susceptible and partially immune populations. We examine the following three scenarios to demonstrate the impact of the weight individuals place on the information about prevalence or severity in the two dynamics, i.e., which information people care about when making a decision to be tested or vaccinated:

#### 1. Scenario 1 (base case)

Susceptible (in primary dynamics) and partially immune (in secondary dynamics) people equally care about both severity and prevalence, given by *α*_1_ = 0.5, *α*_2_ = 0.5 (the base case) for taking a decision to vaccinate or test regardless of their immune status.

#### 2. Scenario 2

Both susceptible and partially immune people prioritize prevalence information, given by *α*_1_ = 0.9, *α*_2_ = 0.9.

#### 3. Scenario 3

Susceptible people prioritize prevalence information and partially immune people prioritize severity, given by *α*_1_ = 0.9, *α*_2_ = 0.1.

The results of scenario 3 (Figure 7 panel c) show that vaccination and testing rates are the greatest, while the vaccination and testing rates of the partially immune population are the lowest. As susceptible populations rapidly become partially immune populations with the model simulations over time, the lower vaccination and testing rates of partially immune populations (second dynamics) resulted in a higher (17%) peak of active cases compared to the base case (scenario 1, Figure 7 panel a). On the other hand, the results of scenario 2 (Figure 7 panel b) show that the vaccination and testing rates of both susceptible and partially immune populations are slightly higher than the base case, as they react more from the prevalence signal to vaccination and testing than base case (90% vs 50%), resulting in a lower (22%) peak of sthe active case than the base case. A shift in prioritization from prevalence to severity information among partially immune individuals (from 90% prevalence and 10% severity to 90% severity and 10% prevalence) while susceptible individuals remain at the same risk perception (90% prevalence and 10% severity) can result in an increase of active cases peak by 50% (from 36 million to 44 million) compared to the case where both (partially immune and susceptible individuals) have higher weight of prevalence than severity (90% prevalence and 10% severity), comparing Figure 7 panel b and c (shift from scenario 2 to scenario 3).

**Figure 7:**
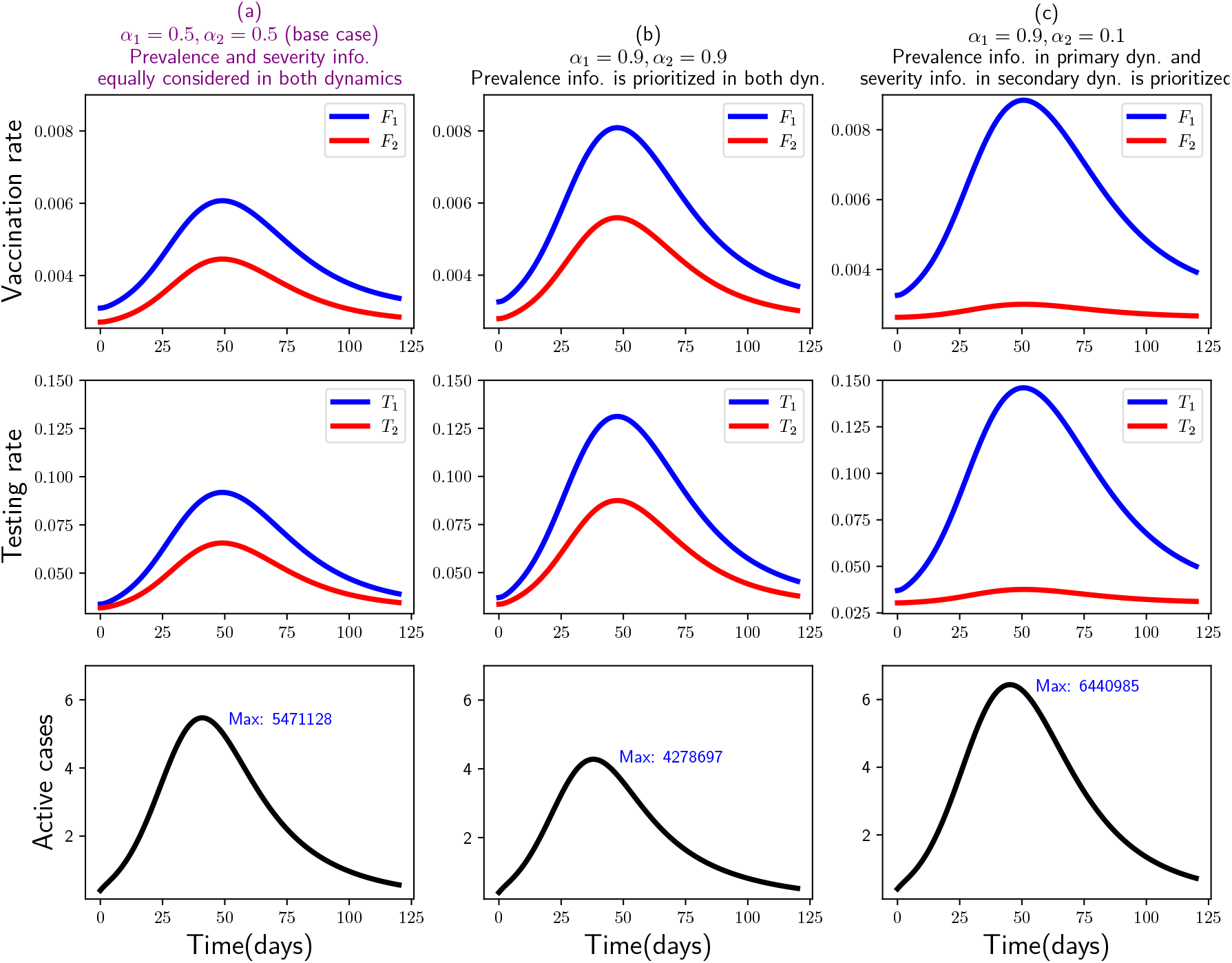
The dynamics of vaccination rate (in primary dynamics *F*_1_, in secondary dynamics *F*_2_), first row, Testing rate (in primary dynamics *T*_1_, in secondary dynamics *T*_2_), second row, and Active cases (*A*_1_ + *A*_2_ + *I*_1_ + *I*_2_), (third row) under three scenarios: first, people in both dynamics equally care about information regarding prevalence and severity, panel (a), second, people in both dynamics care about prevalence than severity information, panel (b), third, people in primary dynamics care about prevalence than severity whereas people in secondary dynamics care about severity than prevalence, panel (c). The parameters *α*_1_ and *α*_2_ indicates the weight given to the prevalence information in primary and secondary dynamics respectively. The remaining (complementary) weights 1 *−α*_1_ and 1 *−α*_2_ are assigned to severity information in the respective dynamics.

We examined the above scenarios based on theoretical assumptions because there is a lack of data regarding how people prioritize information for decisions. The numerical simulation results suggest that differences in risk perceptions among sub-populations may result in different voluntary decisions to vaccination and testing and levels of prevalence may peak consequently. Therefore, the overall epidemic burden is the result of the relative size and distribution of sub-populations and their different risk perceptions and associated care-seeking behaviors. A rise in epidemic can lead to increased voluntary care-seeking behaviors, which can reduce the prevalence. This reduced prevalence can decrease voluntary care-seeking behavior, which in turn can increase the prevalence. Such feedback loop in the model (10) can help us understand a dynamic interplay between disease status and population behaviors. Future work should explore fitting the proposed model to the course of epidemic waves with the seroprevalence and care seeking data from survey, where participants are queried in different time periods of the pandemic about their immune status, risk perception, and contact patterns or care seeking (vaccination and testing) behaviors. Such data can be used to improve the estimates of some of the parameters in the behavior adaptation metrics introduced in this paper.

### (v) Sensitivity of cumulative incidence to parameters

In this subsection we conducted a comprehensive sensitivity analysis using a tornado plot, shown in Figure 8, to show the effect of some selected parameters on the cumulative incidence for a time period of four months (February 01, 2022 – May 31, 2022). The selected parameters are classified as:classical parameters: transmission rate, infectiousness of asymptomatic individuals, proportion of isolated individuals, mandatory vaccination and testing rates, and the behavior related parameters: such as information coverage, information delay, information prioritization, level of reduction in reactivity. For the sensitivity analysis purpose, we set a low and high value for each of these parameters based on the baseline values. The low and high values are fixed to be 50% less than the base values (0.5*×* base value) and 50% higher than the base value (1.5*×* base value), respectively. In Figure 8 the vertical line at the center of the tornado plot represents the cumulative incidence corresponding to the case where all parameter values are fixed at their baseline value, as in Table 1. The effect of the parameters on the cumulative incidence is represented by the length of the bars extending from the base case (center), depending on their low and high values. The findings indicate that, out of all the parameters considered in this analysis, the transmission rate in secondary dynamics, proportion of isolated individuals, and the testing rates are the top drivers of cumulative incidence change, whereas behavioral parameters (such as reactivity to prevalence information, information coverage, reduced reactivity by partially immune people, and prevalence information prioritization by immune people) also have a significant impact. Comparing the sensitivity of parameters in the primary and secondary dynamics, the parameters in the secondary dynamics (e.g., Transmission rate secondary dynamics *β*_2_, prevalence prioritization by partially immune people, *α*_2_) are more sensitive to cumulative incidence than parameters in primary dynamics. This increased sensitivity to parameters in secondary dynamics is due to the large proportion of the initial population in secondary dynamics and the rapid progression from primary to secondary dynamics due to the model formulation.

**Figure 8:**
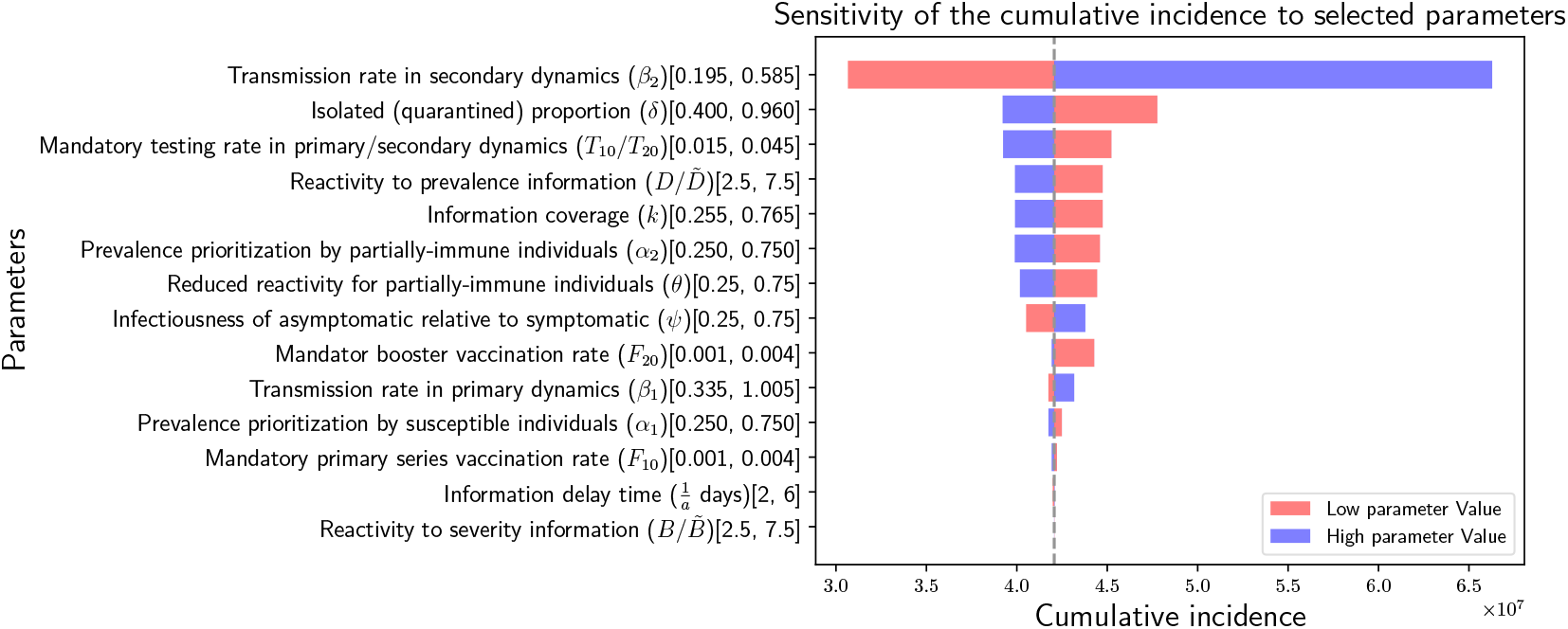
Tornado plot showing the sensitivity of parameters to cumulative incidence over four months (February 01, 2022 - May 31, 2022). The red and blue bars show the cumulative incidence corresponding to a low (0.5*×* base value) and high (1.5*×* base value) values of the parameters, respectively. The center vertical line represents the cumulative incidence when all parameters are fixed at their baseline values. The numbers in the closed bracket for each parameters show the low and high values used for sensitivity analysis: [low, high]. The baseline values for the parameters are as in the Table 1.

## 6 Discussion

In this study, we developed a novel behavior-epidemiology model representing the transmission of COVID-19 that takes into account the behavioral differences among people who are partially immune and those who are not-immune (susceptible) in seeking vaccination and testing [15, 16, 17]. The model outputs were fitted to observed cumulative COVID-19 cases, vaccination and mortality data during the Omicron wave (February 01, 2022 – May 31, 2022) in South Korea. Unlike other behavioral models that employ the information index approach with a single type of information such as disease prevalence [6, 8, 12, 13, 14], our model takes into account that people may have different risk perception (prevalence and severity) and behavior responses (vaccination and testing) across different subgroups by immune status. The overall impact on the prevalence peak in our behavioral model is the result from the non-linear relationship between the number of detected cases that can be information signals to promote voluntary vaccination and testing (decreasing prevalence) and the number of undetected cases that can keep contributing to the transmission (increasing prevalence).

Our stability analysis shows that even the majority (85%) of the population is partially immune, if mandatory vaccination rates decrease below the threshold values (as shown in the yellow frontier line in Figure 4) *R*_*e*_ can be greater than one, and the impact of voluntary behavior parameters become more important. For example, if *R*_*e*_ = 3.17 (obtained by setting *F*_10_ = 10^*−*4^, *F*_20_ = 10^*−*6^, reflecting almost no mandatory booster vaccination (equivalent to 44 people getting mandatory booster vaccination per day, out of the initial population in *S*_2_ and *S*_3_) and 600 daily mandatory primary series vaccination, out of the initial number of people in *S*_1_), the cumulative incidence becomes 43.7 million. This cumulative incidence can be reduced by 20% to 53% by enhancing the voluntary vaccination and testing, which can be achieved by increasing the behavior parameters related to the voluntary vaccination and testing rates from their baseline value– For example, by increasing the level of reactivity of partially immune individuals from the baseline value of 50% to 95% (showing an almost equal level of reactivity as the susceptible individuals), the cumulative incidence can be reduced by 20% (35 million). In addition to this increment, if we increase all other behavioral parameters related to voluntary vaccination and testing rates to some possible maximum value (information coverage (baseline 0.51), prevalence prioritization by susceptible people (baseline 0.5) to 0.95, reactivity to information (baseline 5) to 10) the cumulative incidence can be reduced by 53% (20 million).

Our simulation results indicate that the peak number of active cases can be reduced by up to 16% when partially immune individuals react to information at the same level as susceptible individuals. This is in comparison to a scenario where partially immune individuals have a reduced reactivity to information (50% of the reactivity level of susceptible individuals). This suggests that responsiveness to protective measures by individual immune status can substantially influence the level of future incidence. The result related to information prioritization by susceptible and partially immune individuals showed that over time, if partially immune individuals prioritize severity over prevalence (altering their risk perception), the peak of active cases can increase by up to 17% and 50% compared to when they equally (50% to prevalence and 50% to severity) prioritize both information (base case) and when they prioritize prevalence over severity (90% to prevalence, 10% to severity), respectively. This result shows that when public risk perception changes (e.g., toward severity over prevalence) over the course of the pandemic with multiple variants of epidemic waves, the required target of mandatory vaccination or testing may change, and the public health efforts to control disease may require additional endeavors to reduce the prevalence peak. Furthermore, the variation in prevalence peak based on the risk perception by susceptible and partially immune people underscores the need for further research into how these groups of people perceive diseaserelated information.

The sensitivity analysis result show that the key drivers influencing cumulative incidence are the transmission rate in secondary dynamics and the proportion of isolated (quarantined) individuals among those tested. This demonstrates that non-pharmaceutical interventions such as social distancing and maskwearing remain important to control the spread of the virus besides ongoing testing and vaccination. Moreover, the behavior-related parameters (i.e., reactivity to prevalence information, information coverage, and prevalence information prioritization) substantially contribute to cumulative incidence change, even greater than the mandatory booster vaccination. These results suggest potential disease control mechanisms in two aspects: one is from the government side by increasing information coverage (e.g., by avoiding under reporting, which can be achieved by increasing case detection), and the other is from the population side by an increased level of reactivity and risk perception among partially immune individuals, comparable to that of susceptible people (which enhances the vaccination and testing rates among immune people). In general, the sensitivity results suggest that, when the majority people are initially immune, it is important to consider behavior-related parameters as they can influence total disease burden more significantly than those other standard measures, for example, mandatory vaccination.

There may be various other metrics that could induce behavior changes during a pandemic like COVID-19. For example, authors in a recent study [7] developed a behavioral epidemiology model aiming at assessing the impact of human behavior changes due to factors such as disease-related information received from members of the other group, the level of symptomatic transmission in the community, the proportion of non-symptomatic individuals in the community, the level of publicly available disease-induced mortality information, and fatigue with adherence to control and mitigation interventions in the community, on compliance behavior. One of their results shows that disease-induced mortality has a more significant impact on behavior change compared to the level of symptomatic transmission. This finding somewhat contradicts the results from our model, where the influence of symptomatic infection (disease prevalence) levels is a key driver to the peak of the active cases, rather than the influence of disease severity (measured by the level of deaths and hospitalizations). This discrepancy might be attributed to, for example, the fact that their study examined the impact of each different model output as described above as a signal to change adherence behaviors among the susceptible population in the early stage of the pandemic (thus, most people are susceptible yet) and fit the model to the relatively high mortality rate under the original SARS-CoV-2 virus variant compared to the case during Omicron variants, which resulted in mortality being a driving factor of adherence behavior. In our study, we accounted for behavior adaptation by the individual history of previous exposure and vaccination and explored differing individual reactivity and risk perception/weight by the type of information (between prevalence and severity) and immune status and fitted the model to data during the Omicron epidemic peak (of which the virus variants are characterized by high transmission but low severity [42]). Overall, these findings can be complementary to understand a population behavior dynamics in the early stage of the pandemic, where the adherence behavior of susceptible populations can be a function of various aspects of model outputs, and in the later stage of a pandemic, where care seeking behavior can differ by population immunity status and their risk perception.

Our study has some limitations. First, in the information index approach, there is a lack of enough empiric data to validate the functional forms (in equations (2), (3), (4) and (5)) used for modeling information-dependent vaccination and testing rates. Thus, we took several steps to address this issue: (i)- We iteratively calibrated certain parameters, such as individuals’ reactivity to prevalence or severity information, to ensure that the voluntary vaccination or testing rates reflected the observed data with slower initial growth for the cumulative number of vaccinated and tested individuals, which is typical at the start of a vaccination or testing campaign [30]. (ii)- We also conducted various sensitivity analyses to examine whether the results were robust by changing the input parameter values. Second, our model accounts for the driving forces for vaccination and testing decisions driven by individuals’ risk perceptions about the disease status in the population. However, other factors can influence testing and vaccination decisions, such as access to health care systems, testing procedures, perceptions or beliefs related to vaccine side effects, knowledge of COVID-19 symptoms, etc. [43, 44, 45]. Further more, it is also possible that the individual reactivity to prevalence and severity may differ by other individual characteristics such as age group as well (besides the immune status); in other words, in communities where the majority of people are young, severity information may not be as concerning since COVID-19 deaths predominantly affect older individuals [46]. Conversely, if the majority of the population is old, the average weight of risk perception in the population toward death may be greater than the infection. Indeed, it may not be possible to parameterize all the possible factors and individual heterogeneity that influence willingness to test-ing/vaccination and behaviors in a single model. Moreover, decision-makers are faced with the daunting task of interpreting model predictions while simultaneously estimating how behavioral responses should alter predictions. Despite the given complexities and uncertainties, these estimates might be enhanced by explicitly modeling behavioral responses (i.e., willingness to vaccination as a function of risk and benefit of vaccination) to interventions rather than simply adjusting any assumed constant parameters. Accordingly, our study findings demonstrate the relatively significant impact of behavioral factors on the overall cumulative incidences and thus emphasize the importance of future efforts to collect empiric data and identify driving factors on risk perception and information prioritization among immune and susceptible individuals. Incorporating these behavioral aspects into transmission models may become increasingly useful and important as the epidemic continues and people’s behaviors change.

In summary, this study contributes to the field of epidemiological modeling by illustrating the complex interplay between information, human behavior, and immune status and their impact on disease transmission. To our knowledge, our study is the first attempt to apply an information index with different disease-relevant information (prevalence and severity) among people with different immune statuses and to fit such a model to real-world data. Future studies may further evaluate interplay between various individual status (immunity/age/care seeking history), vaccination, testing and disease transmission, aiming at providing optimal intervention strategies under evolving circumstances.

## Data Availability

All data produced are available online at URL: https://datadryad.org/stash/share/_vm4gFgQ9uxMAFnSbGYLsD41b9l5anwnPKsO6W0YhpU

## Data Accessibility

Numerical simulations were carried out in Python using standard algorithms. We utilized the ‘odeint’ solver from ‘scipy.integrate’ for integrating the model system and employed ‘pyplot’ for figure visualization. All necessary details for reproducing the results are provided in Section 4. The study relies on official COVID-19 data from South Korea, sourced from Our World in Data [30]. The codes and data used are uploaded to Dryad [47]. Use the following links to access them Reviewer URL: https://datadryad.org/stash/share/_vm4gFgQ9uxMAFnSbGYLsD41b9l5anwnPKsO6W0YhpU DOI: https://doi.org/10.5061/dryad.wpzgmsbxp

## Acknowledgements

S.S.S., Y.J. and J.J. gratefully acknowledge the financial support provided by the Ministry of Health & Welfare, Republic of Korea, through the Infectious Disease Medical Safety Project (Grant Number: HG22C0094). B.B. acknowledges the auspices of Italian National Group for Mathematical Physics (GNFM) of the National Institute for Advanced Mathematics (INdAM). B. B. also acknowledges the support of EU funding within the Next Generation EU–MUR PNRR Extended Partnership initiative on Emerging Infectious Diseases (Project No. PE00000007, INF-ACT) and PRIN 2020 project (No. 2020JLWP23) “Integrated Mathematical Approaches to Socio–Epidemiological Dynamics”. We appreciate constructive comments and detailed editorial support from Bernard L. Cook, PhD, at UConn Health.

